# Development of Brain Network Connectivity in Risky Drinkers During Adolescence

**DOI:** 10.1101/2020.10.22.20217992

**Authors:** Simon T. E. Baker, Murat Yücel, Alex Fornito, Andrew Zalesky, Sarah Whittle, Nicholas B. Allen, Dan I. Lubman

**Author notes:** Correspondence concerning this article should be addressed to Simon T. E. Baker, Black Dog Institute, Hospital Road, Randwick, New South Wales 2031, Australia. **Author Note** Simon T. E. Baker is now at Black Dog Institute. Nicholas B. Allen is now at Department of Psychology, University of Oregon.

## Abstract

Alcohol consumption is common in adolescence, a time when the human brain undergoes substantial development, raising concerns about the neurodevelopmental impact of drinking alcohol, especially at high levels. Risky drinking may adversely affect the developing white matter, comprised of axonal fibre pathways that integrate anatomically distributed, and functionally specialised, neural systems. We used diffusion-weighted magnetic resonance imaging (MRI) to perform the first prospective, comprehensive and regionally unbiased connectome-wide analysis of longitudinal changes in inter-regional structural connectivity between 16.5 and 18.8 years of age, comparing adolescents who reported frequent risky drinking (*n* = 20) with those who reported limited risky drinking (*n* = 19) across this developmental period. We found no difference in the development of structural connectivity between these groups, regardless of whether inter-regional connections were quantified using streamline count, fractional anisotropy, mean diffusivity, axial diffusivity, or radial diffusivity. These findings suggest that risky drinking may have limited adverse effects on the development of inter-regional structural connectivity during mid to late adolescence.

---

Alcohol consumption is common in adolescence, with multiple national surveys indicating that up to a half of young people aged around 15–25 report drinking alcohol at levels that are likely to cause significant injury or ill health (i.e., more than four standard drinks on a single occasion) at least monthly (Australian Institute of Health and Welfare, 2011; Johnston, O’Malley, Bachman, Schulenberg, & Miech, 2015; Miech, Johnston, O’Malley, Bachman, & Schulenberg, 2015; Office for National Statistics, 2013). This so-called “risky” or “binge” pattern of alcohol use (hereafter, risky drinking) is a major concern, not only because it is associated with increased morbidity and mortality (e.g., injuries, disabilities, and premature death; Cherpitel et al., 2009; Gore et al., 2011; Rehm, Taylor, & Room, 2006), but also because risky drinking during adolescence is a robust predictor of later alcohol use problems, including alcohol use disorder (AUD; Bonomo, Bowes, Coffey, Carlin, & Patton, 2004; Nelson, Van Ryzin, & Dishion, 2015; Toumbourou et al., 2014; Viner & Taylor, 2007). A further concern is that adolescence is a key developmental period that includes significant structural and functional reorganisation of the brain (Giedd et al., 2015), which raises questions regarding the long-term neurodevelopmental impact of risky drinking (see Conrod & Nikolaou, 2016, for review).

Animal studies (see Spear, 2014, for review) and human magnetic resonance imaging (MRI) studies (see Feldstein Ewing, Sakhardande, & Blakemore, 2014, for review) suggest that the developing brain is particularly sensitive to the effects of alcohol (see Squeglia, Boissoneault, Van Skike, Nixon, & Matthews, 2014, for an integrative review of animal and human studies), with the majority of work focusing on alterations of grey matter. However, a growing body of work suggests that white matter may also be affected by risky drinking during adolescence (see the following reviews: Baker, Yücel, Fornito, Allen, & Lubman, 2013; Elofson, Gongvatana, & Carey, 2013; Monnig, Tonigan, Yeo, Thoma, & McCrady, 2012). Several studies using diffusion-weighted MRI have demonstrated that adolescents who drink more than their peers show microstructural white matter alterations throughout the brain (e.g., Bava, Jacobus, Thayer, & Tapert, 2013; Jacobus, Squeglia, Bava, & Tapert, 2013; Jacobus, Squeglia, Infante, Bava, & Tapert, 2013; Luciana, Collins, Muetzel, & Lim, 2013; see also Baker et al., 2013). Accordingly, there is significant concern that the development of the tightly packed bundles of neuronal axons that constitute a great proportion of white matter tissue may be vulnerable to the damaging effects of alcohol. However, the voxel-based approaches used in prospective and longitudinal studies thus far lack the specificity to pinpoint which particular axonal fibre pathways between anatomically distributed, and functionally specialised, neural systems may be most affected by risky drinking.

To address this gap in the literature, we performed the first prospective, comprehensive and regionally unbiased connectome-wide analysis (Fornito, Zalesky, & Breakspear, 2015; Zalesky, Fornito, & Bullmore, 2010) of longitudinal changes in inter-regional structural connectivity between 16.5 and 18.8 years of age, comparing adolescents who reported frequent risky drinking (*n* = 20) with those who reported limited risky drinking (*n* = 19) across this developmental period. We hypothesised that risky drinkers (i.e., those who consume five or more standard drinks on a single occasion at least monthly, consistent with accepted criteria in the literature [Squeglia et al., 2012, 2015; Squeglia, Spadoni, Infante, Myers, & Tapert, 2009]) would show a distributed pattern of reduced inter-regional structural connectivity, as has been implied by previous diffusion-weighted MRI studies.

## Method

### Participants

Participants were a community sample recruited from schools across metropolitan Melbourne, Australia, as part of a larger longitudinal cohort study (the Orygen Adolescent Development Study) investigating biological, psychological, and social risk factors for psychopathology (see Whittle et al., 2014; Whittle et al., 2008). Our sample included participants who were assessed with MRI during the mid-adolescent (age 16) and late-adolescent (age 18) waves of data collection and screened for the absence of: major medical or neurological condition, medication known to affect nervous system functioning, neurological or incidental radiological abnormality detected as part of routine MRI screening by a hospital radiologist, history of head injury resulting in loss of consciousness, and history of psychotic disorder (schizophrenia or bipolar I). Based on these criteria, and after quality control of MRI data, a sample of 58 adolescents was selected. These adolescents were assigned to one of three groups on the basis of how many times during the 12 months prior to follow-up they engaged in risky drinking, defined as the consumption of five or more standard drinks of alcohol on a single occasion. This classification was based on prior work (Squeglia et al., 2009, 2012, 2015) and was modified according to the distribution of drinking behaviour of adolescents in our sample. Frequent risky drinkers (*n* = 20) were those who engaged in at least 12 risky drinking episodes in the 12 months prior to follow-up. The second group, namely occasional risky drinkers, comprised 19 adolescents who reported between 3 and 11 risky drinking episodes. Adolescents who did not engage in risky drinking (*n* = 9), or who engaged in no more than 2 risky drinking episodes (*n* = 10), were assigned to the comparison group (*n* = 19). Our analyses compared adolescents who reported frequent risky drinking with adolescents who reported limited risky drinking (i.e., the comparison group). Occasional risky drinkers were removed from the analyses in accordance with prior work (Squeglia et al., 2012, 2015).

### MRI Acquisition and Processing

From each participant, we acquired diffusion-weighted and T1-weighted magnetic resonance images on a 3 Tesla Siemens MAGNETOM Trio system located at the Royal Children’s Hospital, Melbourne. Imaging parameters for the high angular resolution diffusion imaging (HARDI) acquisition were as follows: 60 gradient-weighted (b-value = 3,000 s/mm^2^) volumes interspersed with seven T2-weighted (b-value = 0; no gradient weighting) volumes; slice thickness = 2.3 mm; repetition time (TR) = 7,300 msec; echo time (TE) = 104 msec; field of view (FOV) = 240 mm^2^; image matrix = 104 × 104; voxel size = 2.3 mm^3^. For the high-resolution T1-weighted acquisition, imaging parameters were: 176 contiguous 0.9-mm-thick slices; TR = 1,900 msec; TE = 2.24 msec; flip angle = 9°; FOV = 230 mm^2^; image matrix = 256 × 256; voxel size = 0.9 mm^3^. These data were processed using well-validated analysis tools, including FreeSurfer (Fischl, 2012), FMRIB Software Library (FSL; Jenkinson, Beckmann, Behrens, Woolrich, & Smith, 2012), and MRtrix (Tournier, Calamante, & Connelly, 2012), as well as code developed in our laboratory.

### Connectivity Mapping

For each participant, we produced a set of 10 connectivity matrices according to the methods described in Baker et al. (2015). Our pipeline included three major steps. First, we used FreeSurfer (Fischl, 2012) to parcellate the T1-weighted volume into 34 cortical and seven subcortical homologous regions in each hemisphere, totalling 82 distinct brain regions. The cortical regions were identified by spatial normalisation of the cortical surfaces to a probabilistic anatomical atlas (Desikan et al., 2006). The subcortical regions were identified by automatically assigning a unique label to each voxel based on probabilistic information estimated from a manually labelled training set (Fischl et al., 2002). The 68 cortical regions were combined with the 14 subcortical regions to obtain a low-resolution parcellation with 82 non-overlapping regions. We also generated a random parcellation comprising 530 regions (265 per hemisphere) of approximately equal volume (see Zalesky et al., 2010b; Fornito et al., 2011; Fornito, Zalesky, & Bullmore, 2010). This high-resolution parcellation was used to verify our findings. Using FSL (Jenkinson et al., 2012), the inverse transformation matrix from the linear alignment of an average of the seven T2-weighted (b-value = 0) volumes to the T1-weighted volume in Talairach space was applied to both parcellation schemes to align the parcellated regions to native diffusion space (Jenkinson, Bannister, Brady, & Smith, 2002).

In the second step, we used MRtrix (Tournier et al., 2012) to estimate the trajectories of long-range anatomical connections (i.e., axonal fibre pathways). Constrained spherical deconvolution was used to estimate the distribution of fibre orientations at each voxel of the HARDI volume, and then whole brain tractography using a probabilistic fibre-tracking algorithm was performed to obtain a “tractogram” comprising one million streamlines. This approach has been shown to be useful for reconstructing pathways that correspond well with known white matter anatomy (Jeurissen, Leemans, Jones, Tournier, & Sijbers, 2011; Tournier et al., 2012).

In the third step, we combined each parcellation scheme with the tractogram. Specifically, we populated an *N* × *N* adjacency matrix, where *N* is equal to the number of regions (i.e., either 82 or 530), with the number of streamlines intersecting brain region *i* and brain region *j*. A streamline intersecting more than two regions was assumed to connect the pair of regions that were maximally separated according to the Euclidean distance calculated using the centre of gravity of each region. To avoid the inclusion of spurious streamlines crossing the medial longitudinal fissure, interhemispheric streamlines were restricted to those with at least one point of propagation located in a corpus callosum mask. This procedure yielded a connectivity matrix for each parcellation wherein the inter-regional connection weights (or edge weights in graph theory parlance) were quantified using streamline count. To verify our findings, we also quantified inter-regional connection weights using four different scalar measures derived from the eigenvalues of the diffusion tensor that have been proposed to characterise the microstructural properties of white matter, including fractional anisotropy (FA), mean diffusivity (MD), axial diffusivity (AD), and radial diffusivity (RD; Beaulieu, 2009). Connection weights of the FA-weighted connectivity matrices represented the mean FA of connections between brain regions (i.e., FA averaged over all voxels intersected by at least one of the set of streamlines connecting brain regions *i* and *j*). This procedure was repeated for MD, AD, and RD. Thus, we produced 10 different network representations of brain connectivity, including an 82-node and a 530-node streamline count-weighted, FA-weighted, MD-weighted, AD-weighted, and RD-weighted connectivity matrix. We used this multi-measure approach to comprehensively characterise the effects of risky drinking on structural connectivity.

### Statistical Analysis of Connectivity

The general linear model was used to characterise developmental differences in brain network connectivity between adolescents who reported frequent risky drinking and those who reported limited risky drinking during the follow-up period. Separate analyses were conducted for each of the 10 different network representations of brain connectivity. For each analysis, the between-subject covariate in the model was group (risky-drinkers, comparison) and the within-subject covariate was time (mid adolescence [age 16], late adolescence [age 18]). This model was used to test for group-by-time effects at each edge of the connectivity matrix. Connections with a *t*-test statistic exceeding a primary threshold of *p* = .05, two-tailed, uncorrected, were retained for subsequent analysis with the Network Based Statistic (NBS; Zalesky et al., 2010a). The NBS controls the family-wise error (FWE) rate among the multitude of hypotheses tested in a connectome-wide (brain network) analysis. The NBS offers greater statistical power than other methods commonly used to correct for multiple comparisons (e.g., Bonferroni and False Discovery Rate) because statistical inference with the NBS is performed at the level of interconnected subsets of edges, called connected components, rather than at each edge independently. Connected components were assigned a FWE corrected *p*-value using permutation testing (i.e., 10,000 random permutations of the original data). This *p*-value was used to determine whether the size of any observed components significantly exceeded levels expected by chance (FWE corrected, *p* < .05).

## Results

### Demographic and Behavioural Results

Participant characteristics and patterns of alcohol and other drug use are provided in Table 1. Whereas the risky-drinking group was comprised of equal numbers of male and female adolescents (*n* = 10 and *n* = 10, respectively), the comparison group included 14 male and 5 female adolescents; however, this between-group difference was less than conventional statistical significance (*p* = .13). No group differences in age, whole brain volume, or Full Scale Intelligence Quotient (Wechsler, 1999, 2003) were observed between the two groups at the baseline and the follow-up assessment. At both assessments, the risky-drinking group had significantly higher levels of alcohol use across a number of measures of drinking behaviour. The risky-drinking group also had significantly more lifetime cannabis use episodes (range at baseline: 0–3 days; range at follow-up: 0–40 days) than the comparison group (range at follow-up: 0–2 days).

**Table 1.** Participant Characteristics and Patterns of Alcohol and Other Drug Use

|  | Risky-drinking<br>group<br>(*n* = 20) | Comparison<br>group<br>(*n* = 19) | Statistic | *p*-value |
| --- | --- | --- | --- | --- |
| **Stable characteristics** |  |  |  |  |
| Sex | 10 (50%) male;<br>10 (50%) female | 14 (74%) male;<br>5 (26%) female | $\chi^2 = 2.31$ | .13 |
| Handedness | 18 right; 2 left | 19 right; 0 left | $\chi^2 = 2.00$ | .16 |
| **Baseline measures** |  |  |  |  |
| Age in years | 16.48 (0.53) | 16.48 (0.42) | 0.028 | .98 |
| Whole brain volume in mm<sup>3</sup> | 1,255,870<br>(117,061) | 1,241,432<br>(103,228) | 0.41 | .67 |
| Estimated FSIQ (WISC) | 107.40 (13.95) | 109.61 (10.51) | 0.55 | .59 |
| Alcohol use: |  |  |  |  |
| Drinker | 20 (100%) | 10 (53%) | $\chi^2 = 12.32$ | <.001 |
| Lifetime days drinking | 31.00 (28.10) | 6.00 (9.88) | 3.74<sup>a</sup> | .001<sup>a</sup> |
| Days drinking past month | 3.00 (2.79) | 0.16 (0.50) | 4.48<sup>a</sup> | <.001<sup>a</sup> |
| Number of risky drinking<br>episodes past month | 1.53 (1.93) | 0 (0) | 3.46<sup>a</sup> | .003<sup>a</sup> |
| Number of alcohol-related<br>harms past 12 months | 1.25 (1.37) | 0 (0) | 4.08<sup>a</sup> | .001<sup>a</sup> |
| Other drug use: |  |  |  |  |
| Used any tobacco | 9 (45%) | 2 (11%) | $\chi^2 = 5.72$ | .02 |
| Days tobacco use past month | 0.80 (2.67) | 0.05 (0.23) | 1.22 | .23 |
| Used any cannabis | 7 (35%) | 0 (0%) | $\chi^2 = 8.11$ | .004 |
| Lifetime days cannabis use | 0.55 (0.95) | 0 (0) | 2.60<sup>a</sup> | .02<sup>a</sup> |
| **Follow-up measures** |  |  |  |  |
| Age in years | 18.78 (0.36) | 18.86 (0.40) | 0.67 | .51 |
| Whole brain volume in mm<sup>3</sup> | 1,243,162<br>(112,388) | 1,233,855<br>(113,981) | 0.26 | .80 |
| Estimated FSIQ (WASI) | 113.30 (10.88) | 110.32 (10.80) | 0.86 | .40 |
| Alcohol use: |  |  |  |  |
| Drinker | 20 (100%) | 18 (95%) | $\chi^2 = 1.08$ | .30 |
| Lifetime days drinking | 134.20 (85.53) | 16.47 (20.26) | 5.98<sup>a</sup> | <.001<sup>a</sup> |
| Days drinking past 12<br>months | 60.95 (29.82) | 7.26 (6.92) | 7.83<sup>a</sup> | <.001<sup>a</sup> |
| Number of risky drinking<br>episodes past 12 months | 37.50 (21.07) | 0.74 (0.81) | 7.80<sup>a</sup> | <.001<sup>a</sup> |
| Days drinking past month | 6.85 (4.49) | 1.74 (2.10) | 4.60<sup>a</sup> | <.001<sup>a</sup> |
| Number of risky drinking episodes past month | 3.55 (2.80) | 0.21 (0.54) | 5.23<sup>a</sup> | <.001<sup>a</sup> |
| Number of alcohol-related harms past 12 months | 2.55 (1.79) | 0.16 (0.50) | 5.74<sup>a</sup> | <.001<sup>a</sup> |
| Other drug use: |  |  |  |  |
| Used any tobacco | 13 (65%) | 4 (21%) | $\chi^2 = 7.65$ | .006 |
| Days tobacco use past month | 7.30 (12.14) | 0.16 (0.69) | 2.63<sup>a</sup> | .02<sup>a</sup> |
| Used any cannabis | 12 (60%) | 2 (11%) | $\chi^2 = 10.36$ | .001 |
| Lifetime days cannabis use | 7.50 (10.36) | 0.16 (0.50) | 3.17<sup>a</sup> | .005 |

Values are mean (*SD*), unless otherwise indicated. Statistics are independent-samples *t*-test
statistics, unless otherwise indicated. All *p*-values are two-tailed. <sup>a</sup>Adjusted test statistic and adjusted *p*-value reported because the assumption of homogeneity of variances was violated.
FSIQ = Full Scale Intelligence Quotient; WASI = Wechsler Abbreviated Scale of Intelligence (Wechsler, 1999); WISC = Wechsler Intelligence Scale for Children—4th Edition (Wechsler, 2003).

### Connectivity Results

Using the 82-node anatomical parcellation, the group average connection density, *κ*, for adolescents who reported frequent risky drinking was 75% ± 3.5% *SD* at baseline and 73% ± 3% *SD* at follow-up, and for adolescents who reported limited risky drinking the group average *κ* was 75% ± 4% *SD* at baseline and 74% ± 3% *SD* at follow-up. With the 530-node random parcellation, the group average *κ* for the risky-drinking group was 24% ± 1.5% *SD* at baseline and 23% ± 1% *SD* at follow-up, and for the comparison group the group average *κ* was 24% ± 1.5% *SD* at baseline and 24% ± 1% *SD* at follow-up. Thus, no between-group differences were found in the overall number of inter-regional connections at either time point using either parcellation resolution (see Table 2).

**Table 2.** Connection Density

|  | Risky-drinking<br>group | Comparison<br>group | *t* statistic | *p*-value |
| --- | --- | --- | --- | --- |
| **82-node parcellation** |  |  |  |  |
| Baseline | 75 (3.5) | 75 (4) | 0.23 | .82 |
| Follow-up | 73 (3) | 74 (3) | 0.62 | .54 |
| **530-node parcellation** |  |  |  |  |
| Baseline | 24 (1.5) | 24 (1.5) | 0.14 | .89 |
| Follow-up | 23 (1) | 24 (1) | 1.88 | .07 |

Values are mean (*SD*) connection density, $\kappa$ , calculated as the proportion (%) of all possible connections actually present. Statistics are independent-samples *t*-test statistics. All *p*-values are two-tailed.

We used the NBS to identify subnetworks comprised of structural connections showing developmental differences between the risky-drinking group and the comparison group. Using a two-tailed *t*-test statistic threshold of *p* = .05 and a component-wide FWE corrected threshold of *p* = .05, there were no significant group-by-time effects at either parcellation resolution using either streamline count or any of the four tensor-based measures (i.e., FA, MD, AD, and RD) as connectivity weight. Relaxing the primary threshold to *p* = .10, two-tailed, yielded the same results.

## Discussion

This is the first study to use a longitudinal design to assess whether the development of axonal fibre pathways between pairs of brain regions is affected by risky drinking between mid and late adolescence. Contrary to expectations, we found no difference in the development of long-range inter-regional structural connectivity between adolescents who reported frequent risky drinking and those who reported limited risky drinking during this period. These findings suggest that the impact of risky drinking on mid-to late-adolescent brain development does not manifest as a major deviation from normal developmental changes in inter-regional structural connectivity, as measured with diffusion-weighted MRI connectomics.

Previous longitudinal MRI studies have suggested that risky drinking in adolescence may be detrimental to white matter development. These include three diffusion-weighted MRI studies that reported microstructural white matter alterations throughout the brain in adolescents who either initiated (Jacobus et al., 2013b) or continued (Bava et al., 2013; Jacobus et al., 2013a) heavy alcohol and cannabis use between mid and late adolescence. A fourth study followed adolescents between the ages of 14–19 (range at baseline) and 16–22 (range at follow-up) years and found evidence of microstructural alterations in those who started drinking alcohol (Luciana et al., 2013). Together with our data, these findings suggest that risky drinking in adolescence appears to be associated primarily with small-scale (perhaps subtle) white matter alterations, rather than disturbances of large-scale inter-regional structural connectivity. It is thus possible that in the early stages of risky drinking, white matter disruptions are focal, and are not extensive enough to be detected in comparisons of tract-averaged measures of tissue microstructure, as studied here. In other words, the scale of analysis (voxel-based vs. connectome-wide) may explain the discrepancy in findings. It remains to be seen whether these microstructural alterations progress into more extensive structural connectivity disturbances later on, especially if risky drinking continues or escalates.

*In vivo* MRI and post-mortem studies of people with chronic AUD have found evidence of white matter pathology (Monnig et al., 2012; Sullivan & Pfefferbaum, 2005), and a recent study (Kuceyeski, Meyerhoff, Durazzo, & Raj, 2013) found reduced structural connectivity among grey matter regions belonging to the brain’s reward system. Such findings suggest that gross (macroscopic) white matter changes may become evident with long-term heavy drinking and/or progression to chronic illness. It will be important for future studies to examine when in the course of heavy drinking and/or AUD gross white matter changes appear. An important question, however, is the extent to which these changes might reflect the direct effect of alcohol or the compounding effects of genetic, behavioural, social, environmental, nutritional, and premorbid factors. These cofactors therefore need to be considered in any rigorous study of the neurobiological sequelae of alcohol use.

Our findings should be interpreted in the light of some important considerations. The adolescent sample studied here was representative of the community from which it was drawn. As a result, levels of alcohol use were generally low, with few adolescents experiencing multiple or relatively severe alcohol-related harms. This limited range in alcohol outcomes may have reduced statistical power to detect statistically significant effects when comparing groups. Conversely, it provides a more accurate representation of drinking behaviour in a non-clinical adolescent population.

The adolescents in our study reported relatively limited cannabis use compared to the adolescents in most previous studies (e.g., Bava et al., 2013; Jacobus et al., 2013a, 2013b). Interestingly, one of these studies found microstructural alterations in adolescents who transitioned from limited prior alcohol and cannabis use (age range: 16–18 years) to heavy drinking and heavy cannabis use (age range: 19–22 years) in the absence of any microstructural alterations in adolescents who initiated heavy drinking only (Jacobus et al., 2013b). Thus, further work is required to disentangle the unique effects of alcohol and other drugs such as cannabis on white matter development.

Several limitations of our study should also be noted. First, our sample size was small but adequate compared to previous studies that found microstructural white matter alterations (group *n*s range from 8 to 41 alcohol-or substance-using adolescents and 16 to 51 adolescents who have had no or limited exposure to alcohol or other drugs). Our sample size also precluded an analysis of sex differences, which are known to influence brain development (Giedd et al., 2015; Ingalhalikar et al., 2014) and the effects of alcohol (Becker, McClellan, & Reed, 2017). Second, participants had already started drinking alcohol—and, in some cases, using tobacco and cannabis—prior to the baseline MRI assessment in mid adolescence. However, we examined the development of structural connectivity *within* individuals; that is, our analysis focused on patterns of change relative to each individual’s baseline. Third, the fact that occasional risky drinkers were removed from the analyses to increase the likelihood of finding alcohol-induced developmental differences in brain network connectivity between frequent risky-drinkers and the comparison group means that our study did not utilise all available data. Finally, although the longitudinal design of this study is a strength, the follow-up period was limited to approximately two years and did not include emerging adulthood when further escalation and entrenchment of drinking behaviour occurs. Studying the evolution of brain changes as risky drinking continues or escalates during this developmental period will be an important goal for future research.

The findings reported here suggest that risky drinking may have limited adverse effects on the development of inter-regional structural connectivity between mid and late adolescence. While previous studies examining brain development and alcohol use in adolescence support the notion that risky drinking may be detrimental to white matter, that impact may be confined to microstructural alterations localised to specific areas of white matter, which may not be detected in connectome-wide analyses. Determining when in the course of risky/heavy drinking and/or AUD these microstructural alterations deteriorate to more profound structural connectivity disturbances will be an important area of future research.

## Data Availability

Data may be available on request

